# An Integrative Multimodal Model for Early Diagnosis of Dementia and Differential Diagnosis of Alzheimer’s Disease Using Neuroimaging, Polygenic Risk, and Cognitive Assessments

**DOI:** 10.64898/2026.07.31.26359317

**Authors:** Tahir Tekin Filiz, Vera Fominykh, Karin Persson, Mona Michelet, Iris J. Broce, Ingrid Tøndel Medbøen, Stina Aam, Alexey Shadrin, Dag Alnæs, Lavinia Athanasiu, Xin Wang, Gisele Sanda, Ingvild Tina Saltvedt, Anne-Brita Knapskog, Geir Selbæk, Anders M. Dale, Ole A. Andreassen, Oleksandr Frei

## Abstract

**Background:** Early diagnosis and etiological classification of dementia remain challenging, as clinicians typically lack tools to integrate cognitive, neuroimaging, and genetic data quantitatively. We developed and validated multimodal risk models to support early diagnosis of dementia and differential diagnosis of Alzheimer’s disease (AD) versus non-AD dementias in real-world clinical settings and translated model outputs into individualized risk reports.

**Methods:** Utilizing real-world clinical cohorts (n = 1,100 for early diagnosis of dementia, using clinical diagnoses up to three years after clinical assessment; n = 788 for AD differential diagnosis) from Norwegian Memory Clinics, we trained and validated the Multimodal Hazard Score for Real-World Data (MHS-RWD) model integrating demographics (age, sex), cognitive assessments (MMSE-NR3 or CERAD 10-word delayed recall), the MRI-derived Imaging Hazard Score, and the Polygenic Hazard Score. Discrimination performance was examined using the area under the receiver operating characteristic curve (AUC).

**Results:** In real-world clinical data, the MHS-RWD consistently outperformed any single predictor used alone. For early diagnosis of dementia, the full model achieved an AUC of 0.89 in females and 0.84 in males. For the differential diagnosis of AD from other dementias, the multimodal model yielded an AUC of 0.91 in females and 0.83 in males. A patient-level risk report was designed to present individualized risk estimates.

**Conclusions:** Multimodal integration of cognitive, neuroimaging, and polygenic data in the MHS-RWD tool yields strong discrimination for both early diagnosis of dementia and AD differential diagnosis. The tool relies on data obtainable in clinical care, and genetic information that is becoming increasingly available in routine practice. Delivered through intuitive patient-level risk reports, it could support etiologically informed dementia decisions in real-world settings, with potential utility in primary care.

## Introduction

Dementia is a growing global health challenge, with profound personal, social, and economic consequences driven largely by aging populations and increasing life expectancy [1–4]. At the healthcare system level, this rising burden is compounded by limited specialist capacity and the complexity of early, etiologically accurate diagnosis, creating a need for scalable and reliable approaches to dementia management [5,6]. Across healthcare systems, primary care often serves as the first point of contact for cognitive symptoms, and in several countries national guidelines recommend or permit that typical, uncomplicated cases of dementia be diagnosed in primary care [7–10]. This places responsibility on general practitioners (GPs) not only to recognize cognitive impairment but also, where appropriate, to assess underlying etiology, at minimum distinguishing Alzheimer’s disease (AD) from non-AD dementias such as vascular dementia, dementia with Lewy bodies, or frontotemporal dementia [8,10]. While structural brain MRI is commonly included in dementia work-up, primarily to exclude alternative causes of cognitive impairment, such as stroke or mass lesions, GPs typically lack the specialist training needed to extract and interpret quantitative markers of neurodegeneration from these scans [6,11]. In parallel, advances in polygenic risk scoring enable estimation of an individual’s inherited liability for AD, while cognitive assessments provide functional evidence of impairment. However, these sources of information are currently not combined in a systematic and accessible way in clinical routine. Thus, there is a need for clinical decision support tools that integrate automated quantitative analysis of clinically acquired MRI with genetic risk information, and cognitive assessments [11]. With disease-modifying AD therapies now emerging, such tools are becoming increasingly relevant for early diagnosis and for identifying individuals on a trajectory toward AD. Primary care is uniquely positioned to meet this challenge if equipped with the right tools.

Previous work has progressively advanced individualized, time-to-event risk modeling for AD by integrating biomarkers across modalities. McEvoy et al. introduced the Imaging Hazard Score (IHS) and demonstrated that regional MRI-derived measures of brain atrophy, both cross-sectional and longitudinal, can be combined to stratify individuals with mild cognitive impairment (MCI) by their risk of conversion to AD dementia within a survival-analysis framework [12]. Building on this concept, Desikan et al. developed the Polygenic Hazard Score (PHS), which leveraged multiple AD-associated genetic variants, beyond *APOE*, to estimate age-specific genetic risk for disease onset and progression [13]. The PHS linked genetic liability to longitudinal clinical decline, neuroimaging markers, and neuropathology, and showed that the polygenic information can effectively differentiate individual risk trajectories. More recently, Reas et al. introduced the Multimodal Hazard Score (MHS), integrating genetic risk, neuroimaging measures, and memory performance to predict time to dementia onset among individuals with MCI or subjective cognitive impairment (SCI) [14]. Together, these studies established hazard-based, multimodal modeling as a powerful framework for personalized risk prediction in AD.

However, prediction models developed in research cohorts may not retain the same performance in heterogeneous real-world clinical populations [15,16]. In contrast to controlled research studies, clinicians in real-world settings face greater diagnostic uncertainty, particularly at the earliest stages of cognitive decline. Moreover, much of the previous hazard-based work in AD has focused on predicting progression from well-defined clinical states such as MCI or established dementia, implicitly assuming that the current disease stage has already been accurately determined [12–14,17–19]. In real-world healthcare, this initial differentiation between normal aging, MCI, and dementia is itself a core challenge, especially in the early disease phases, and precisely where clinical decision support is most needed.

In the current study, we extended prior hazard-based modeling by improving the MHS introduced by Reas et al. [14] to MHS-RWD for use in real-world healthcare settings and validated its performance for early diagnosis of dementia. We then developed and validated the MHS-RWD for differential diagnosis of AD versus non-AD dementias by incorporating a focused memory measure using data from the Norwegian Registry of Persons Assessed for Cognitive Symptoms (NorCog) [20]. Finally, we translated these predictive models into practice by designing a patient-level risk report that presents individualized risk estimates in an intuitive, clinically interpretable format tailored to the workflow and clinical decision-making needs of real-world clinicians.

## Methods

### Cohort description

Data were drawn from the NorCog, including patients from the Oslo University Hospital (OUS) Memory Clinic (Oslo, Norway) and the geriatric outpatient clinic at St. Olavs Hospital (Trondheim, Norway). The following variables were extracted at study baseline: demographics (sex, age, site); cognitive measures (Mini-Mental State Examination Norwegian Revision, MMSE-NR3 [21] and Consortium to Establish a Registry for Alzheimer’s Disease (CERAD) neuropsychological battery 10-word delayed recall [22]); and clinical diagnosis data, including overall cognitive impairment stage (SCI, MCI, or dementia), Clinical Dementia Rating (CDR) score [23], MCI etiological classification (likely due to AD, likely due to another dementia, likely not due to dementia, unclear cause), and dementia etiology codes (early-onset AD, late-onset AD, mixed AD, vascular and other specific dementias including frontotemporal dementia, dementia with Lewy bodies, and Parkinson’s disease dementia, unspecified dementia), and other primary diagnosis codes. Under the NorCog protocol, these diagnosis codes were assigned by specialists after completion of the baseline clinical evaluation and interdisciplinary consensus review. Diagnoses were established according to ICD-10 alongside optional etiology-specific research criteria [20].

For OUS participants classified as MCI or dementia during the baseline assessment, we applied the most recent follow-up code recorded within three years to achieve a more accurate etiological classification. Follow-up ICD-10 codes were extracted from the OUS hospital electronic health record system and converted to NorCog diagnostic codes using the standard registry procedure.

### Classification outcome definitions

For model development, we defined two classification outcomes, with two separate definitions of cases and controls. For both outcomes, predictors were taken from the baseline assessment, while diagnostic information from baseline assessment and up to three years of follow-up was used to define outcome status. We excluded individuals aged 55 or younger because the IHS component was based on the Alzheimer’s Disease Neuroimaging Initiative (ADNI) data, which recruits individuals older than 55 [24]. We also excluded those with no cognitive impairment or another primary non-dementia diagnosis.

For the early diagnosis of dementia, cases were defined as individuals diagnosed with dementia during the baseline assessment, those with any recorded dementia etiology within three years after baseline, or those classified as MCI with a subclass suggestive of prodromal dementia (either likely due to AD or likely due to another dementia). Controls were patients with subjective complaints or mild impairment who had neither a dementia code nor an MCI subclass consistent with an underlying dementia disease. By combining diagnoses at baseline assessment with diagnoses recorded within three years of follow-up, this outcome captures current or near-future clinical identification of dementia rather than incident dementia alone. Here, early diagnosis refers to the timing of detection within the diagnostic pathway and the tool’s intended use, not to clinical stage (mild vs severe dementia) or to the underlying biological progression of the disease.

For the differential diagnosis of AD outcome, the cohort was restricted to subjects in the MCI or dementia stages, and subjects with unspecified dementia etiology were excluded. Cases were defined as individuals whose etiology codes point to AD, or patients at MCI stage marked as likely due to AD. Controls were the remaining MCI/dementia patients whose etiology points to non-AD causes.

A flowchart of cohort selection and outcome-specific analytic samples for the full MHS-RWD models is provided in Supplementary Figure 3.

### Imaging hazard scores and polygenic hazard scores for MHS-RWD

For computation of IHS, MRI scans were retrieved from the clinical PACS system, then T1 MPRAGE-like sequences were identified from DICOM metadata, and each scan was segmented with Cortechs.ai NeuroQuant; we then applied the imaging hazard model by Reas et al. [14] previously trained on ADNI subjects to those segmentations to derive IHS values for NorCog participants, restricting to examinations obtained within six months before or after baseline so imaging and clinical phenotyping remained aligned.

For computation of PHS, the polygenic hazard model originally trained in the EADB study by Akdeniz et al. was applied to NorCog’s genotype data [19]. Samples were genotyped at deCODE Genetics and imputed as previously described [19]. Individual PHS scores were computed using PLINK (--score function), ensuring compatibility between the published weights and the imputed variant set [25,26].

### MHS-RWD Model Specification, Training, and Validation

For the early diagnosis of dementia, we trained a multivariable model including age, sex, MMSE-NR3, IHS, and PHS predictors. For the AD differential diagnosis model, MMSE-NR3 was replaced with the CERAD 10-word delayed recall test to better capture the memory domain.

Model weights for both outcomes were estimated in the St. Olavs Hospital cohort using unregularized logistic regression. Prior to fitting, all quantitative predictors except age and PHS were pre-residualized on age, and all model inputs were then standardized to zero mean and unit variance using training-set moments, with age-residualization and standardization parameters learned in the training data and then carried forward unchanged to avoid leakage. For comparison, the baseline model was trained using age and sex only.

Validation in OUS Memory Clinic data was carried out for the full and baseline models, as well as for all individual predictors and the score of the MHS model previously fitted on ADNI data by Reas et al. [14]. The validation was carried out in a sex-stratified manner to reflect established sex-related differences in dementia and AD prevalence, cognitive trajectories, brain atrophy patterns, and biomarker expression, which may also affect model calibration and discrimination if sexes are pooled [27,28].

Model discrimination was summarized by the area under the ROC curve (AUC) together with the ROC coordinates (false-positive rate, FPR; true-positive rate, TPR) across score thresholds. For clinical-style reporting, we additionally defined three score bands using cut points set at the 20th and 60th percentiles of the score distribution in the OUS memory-clinic data, corresponding to a negative test result, an inconclusive (grey zone) result, and a positive test result. Within these bands, we estimated the observed outcome prevalence (risk) in the positive, inconclusive, and negative groups. We then reported band-based operating characteristics: coverage, defined as the fraction of individuals receiving a definitive result (positive or negative, i.e., not inconclusive); and conditional performance metrics (excluding the inconclusive group), including conditional sensitivity and conditional specificity, each calculated only among those classified as positive or negative. Confidence intervals at 95% for all readouts were computed using bootstrap resampling with replacement within the validation cohort.

### Individual patient reports

For each outcome, individualized patient reports were constructed to visualize the distribution of model-derived risk scores relative to the reference dataset. Each report consists of one main panel displaying the combined model score and three component panels showing the cognitive test, imaging, and genetic scores as smoothed density curves estimated using kernel density estimation (KDE), weighted by group prevalence, displayed separately for cases and controls. Cognitive and imaging component scores are adjusted for age so they can be interpreted as age-specific deviations from the expected level, with higher values indicating greater abnormality or impairment, whereas the genetic score reflects inherited risk. Score regions are defined using the same percentile-based cut points (the 20th and 60th percentiles). At the patient’s score, group membership probabilities were estimated from the corresponding density curves and clipped to “10% or less” and “90% or more” at the extremes.

## Results

### Cohort demographics

The dataset used for model training and validation was restricted to participants with complete data for all predictors, with sex-stratified validation sub-cohorts differing by outcome. For early diagnosis of dementia, the male subset included 203 cases and 150 controls, whereas the female subset comprised 189 cases and 103 controls. For AD differential diagnosis, the male validation cohort contained 100 AD cases and 119 non-AD controls, while the female cohort included 119 AD cases and 74 controls. Demographic and clinical characteristics of the training and validation cohorts are summarized in Table 1 for the early diagnosis of dementia model and in Table 2 for the AD differential diagnosis model. Among participants with a recorded global CDR score, the large majority were at the lower end of the severity spectrum (CDR ≤ 1), with only a small number at CDR 2 and 3 (Supplementary Table 2).

**Table 1.** Cohort characteristics for the early diagnosis of dementia cohort, stratified by data split (training/validation), group (cases/controls), and sex. Values are mean (standard deviation). MMSE-NR3: Mini-Mental State Examination Norwegian Revision; IHS: Imaging Hazard Score; PHS: Polygenic Hazard Score; N: sample size.

| Split | Group | Sex | N | Age | MMSE-NR3 | IHS | PHS |
| --- | --- | --- | --- | --- | --- | --- | --- |
| Train | Cases | Male | 168 | 76.77 (6.33) | 21.47 (4.79) | 1.57 (1.33) | -0.73 (0.52) |
| Train | Cases | Female | 181 | 76.92 (6.08) | 20.85 (4.24) | 1.46 (1.19) | -0.65 (0.47) |
| Train | Controls | Male | 54 | 76.39 (5.38) | 25.30 (3.64) | 0.67 (1.23) | -0.84 (0.42) |
| Train | Controls | Female | 52 | 76.29 (6.29) | 25.04 (3.43) | 0.77 (1.22) | -0.75 (0.53) |
| Validation | Cases | Male | 203 | 70.96 (7.17) | 24.72 (4.10) | 0.74 (1.41) | -0.68 (0.56) |
| Validation | Cases | Female | 189 | 69.90 (7.27) | 23.54 (4.23) | 0.66 (1.24) | -0.56 (0.52) |
| Validation | Controls | Male | 150 | 67.58 (7.75) | 27.88 (2.35) | -0.86 (1.21) | -0.94 (0.43) |
| Validation | Controls | Female | 103 | 66.16 (7.53) | 28.05 (2.38) | -1.03 (1.12) | -0.93 (0.44) |

**Table 2.**
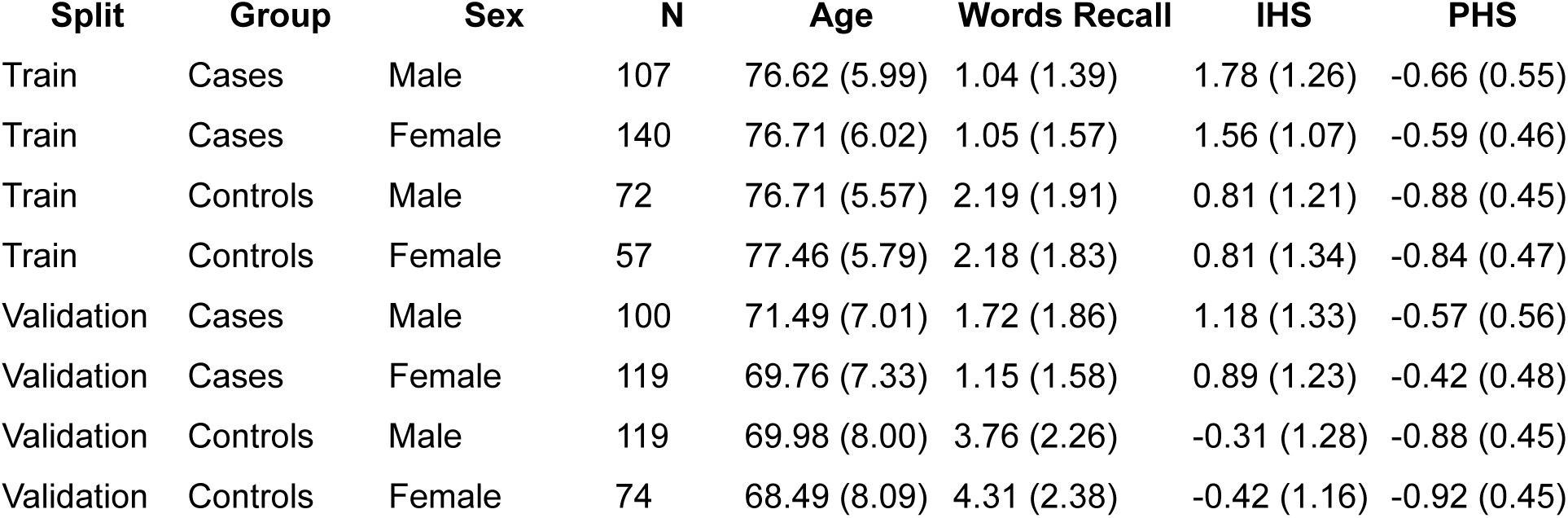
Cohort characteristics for the Alzheimer’s disease differential diagnosis cohort, stratified by data split (training/validation), group (cases/controls), and sex. Values are mean (standard deviation). Words Recall: CERAD 10-word delayed recall; IHS: Imaging Hazard Score; PHS: Polygenic Hazard Score; N: sample size.

| Split | Group | Sex | N | Age | Words Recall | IHS | PHS |
| --- | --- | --- | --- | --- | --- | --- | --- |
| Train | Cases | Male | 107 | 76.62 (5.99) | 1.04 (1.39) | 1.78 (1.26) | -0.66 (0.55) |
| Train | Cases | Female | 140 | 76.71 (6.02) | 1.05 (1.57) | 1.56 (1.07) | -0.59 (0.46) |
| Train | Controls | Male | 72 | 76.71 (5.57) | 2.19 (1.91) | 0.81 (1.21) | -0.88 (0.45) |
| Train | Controls | Female | 57 | 77.46 (5.79) | 2.18 (1.83) | 0.81 (1.34) | -0.84 (0.47) |
| Validation | Cases | Male | 100 | 71.49 (7.01) | 1.72 (1.86) | 1.18 (1.33) | -0.57 (0.56) |
| Validation | Cases | Female | 119 | 69.76 (7.33) | 1.15 (1.58) | 0.89 (1.23) | -0.42 (0.48) |
| Validation | Controls | Male | 119 | 69.98 (8.00) | 3.76 (2.26) | -0.31 (1.28) | -0.88 (0.45) |
| Validation | Controls | Female | 74 | 68.49 (8.09) | 4.31 (2.38) | -0.42 (1.16) | -0.92 (0.45) |

### Model performance

For both outcomes, multimodal integration consistently yielded the strongest discrimination (Table 3). For early diagnosis of dementia, the fully integrated MHS-RWD model combining age, MMSE-NR3, imaging hazard score (IHS), and polygenic hazard score (PHS) achieved the highest discrimination and performed nearly identically to the MHS trained in ADNI (ADNI-MHS), with AUCs of approximately 0.89 in females and 0.84 in males (Figure 1A), indicating strong cross-cohort consistency despite independent model development. More broadly, the MHS-RWD multimodal model outperformed all individual domains, including cognitive, imaging, and genetic predictors considered separately, confirming the complementary information captured by multimodal risk integration.

**Fig 1.**
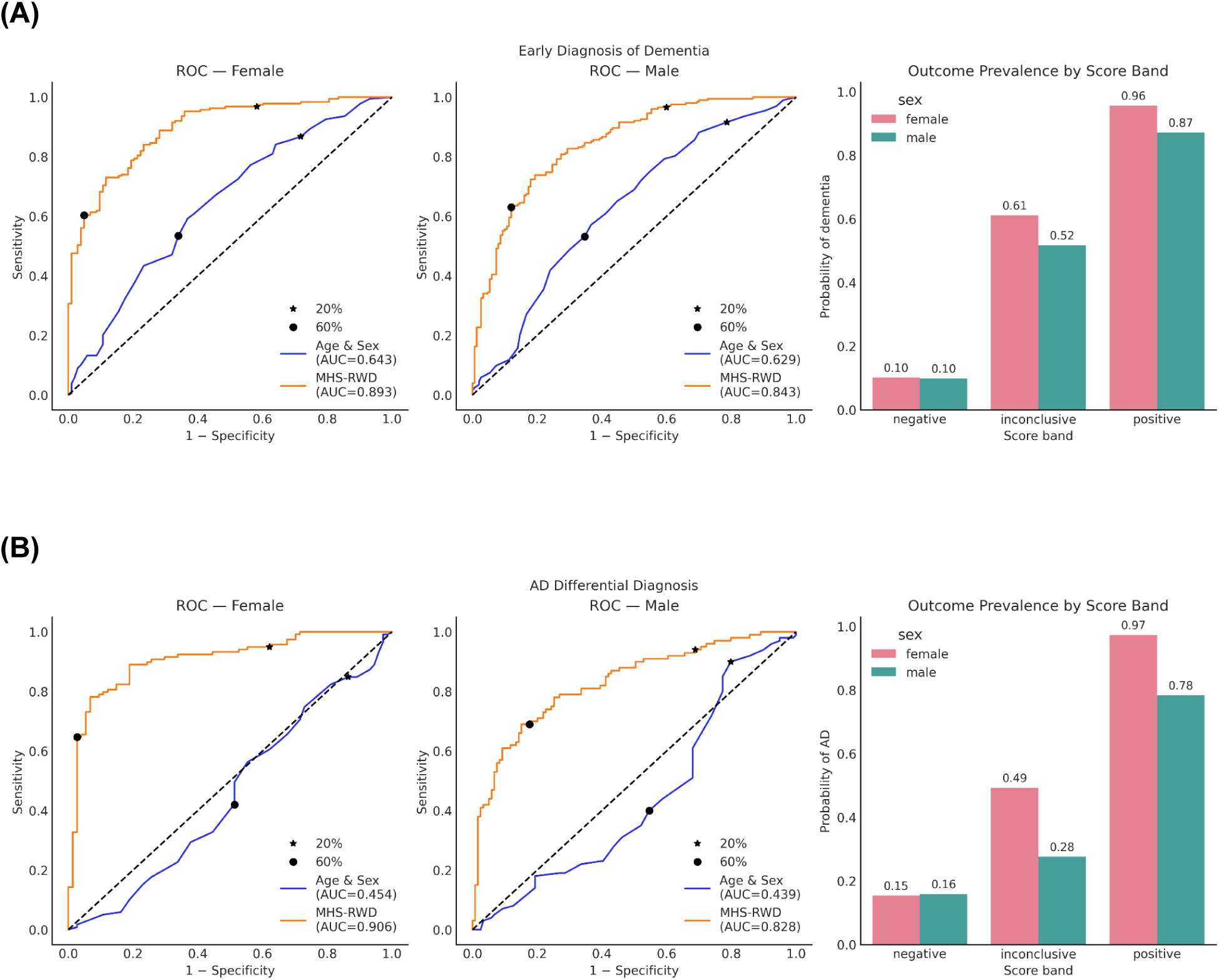
Accuracy of the models for early diagnosis of dementia and differential diagnosis. The figure presents sex-stratified model performance and risk stratification for the two outcomes. Panel (A) shows results for early diagnosis of dementia; panel (B) shows results for Alzheimer’s disease differential diagnosis. In each panel, receiver operating characteristic (ROC) curves are displayed separately for females and males, comparing a baseline age-only model with MHS-RWD; the corresponding area under the curve (AUC) values are reported in the legends. Star and dot markers on the ROC curves indicate predefined percentile-based thresholds used to define negative, inconclusive (grey zone), and positive score regions. To the right of each ROC pair, bar plots summarize the sex-stratified probability of the clinical outcome within these three score categories.

**Table 3.** Sex-stratified model performance for early diagnosis of dementia and differential diagnosis of Alzheimer’s disease, reported as area under the receiver operating characteristic curve (AUC). Models include age, Mini-Mental State Examination Norwegian revision (MMSE-NR3), imaging hazard score (IHS), polygenic hazard score (PHS), multimodal hazard score trained on ADNI (ADNI-MHS), and CERAD 10-word delayed recall (Words Recall). 95% confidence intervals are denoted in parentheses.

| **Outcome** | **Model** | **AUC – female** | **AUC – male** |
| --- | --- | --- | --- |
| **Early diagnosis of dementia** | Age | 0.643 (0.578-0.708) | 0.629 (0.567-0.689) |
|  | Age+MMSE-NR3+IHS | 0.882 (0.841-0.918) | 0.832 (0.787-0.873) |
|  | Age+MMSE-NR3+IHS+PHS | 0.893 (0.855-0.929) | 0.843 (0.798-0.883) |
|  | IHS | 0.847 (0.805-0.889) | 0.805 (0.759-0.847) |
|  | ADNI-MHS | 0.892 (0.854-0.928) | 0.840 (0.798-0.880) |
|  | MMSE-NR3 | 0.829 (0.777-0.876) | 0.749 (0.696-0.798) |
|  | PHS | 0.696 (0.632-0.756) | 0.631 (0.573-0.688) |
| **Differential diagnosis (AD)** | Age | 0.454 (0.370-0.544) | 0.439 (0.362-0.519) |
|  | Age+Words Recall | 0.870 (0.815-0.920) | 0.743 (0.675-0.807) |
|  | Age+Words Recall+IHS | 0.875 (0.823-0.921) | 0.804 (0.743-0.856) |
|  | Age+Words Recall+IHS+PHS | 0.906 (0.860-0.947) | 0.828 (0.771-0.879) |
|  | IHS | 0.779 (0.710-0.843) | 0.795 (0.730-0.851) |
|  | MMSE-NR3 | 0.730 (0.652-0.799) | 0.663 (0.589-0.733) |
|  | PHS | 0.763 (0.690-0.831) | 0.675 (0.598-0.747) |
|  | Words Recall | 0.870 (0.814-0.921) | 0.748 (0.680-0.811) |

**Table 4.** Sex-stratified discrimination and risk by score band. Sex-stratified discrimination performance and score-band risk estimates for models developed for early diagnosis of dementia and differential diagnosis of Alzheimer’s disease in the reference cohort. Sensitivity and specificity are reported conditional on receiving a definitive classification (i.e., excluding the inconclusive score band). Risk estimates correspond to the observed outcome probability within the positive and negative score bands. 95% confidence intervals are denoted in parentheses. Overall coverage of definitive (non-inconclusive) classifications was approximately 60% across sex and outcome strata.

| **Metric** | **Male – Early diagnosis of dementia** | **Male – Differential diagnosis (AD)** | **Female – Early diagnosis of dementia** | **Female – Differential diagnosis (AD)** |
| --- | --- | --- | --- | --- |
| Sensitivity (conditional) | 0.946 (0.90, 0.98) | 0.908 (0.84, 0.97) | 0.949 (0.91, 0.98) | 0.926 (0.86, 0.98) |
| Specificity (conditional) | 0.780 (0.68, 0.87) | 0.661 (0.53, 0.78) | 0.914 (0.84, 0.98) | 0.943 (0.86, 1.00) |
| AUC | 0.843 (0.80, 0.88) | 0.828 (0.77, 0.88) | 0.893 (0.85, 0.93) | 0.906 (0.86, 0.95) |
| Risk (positive) | 0.872 (0.82, 0.93) | 0.784 (0.70, 0.87) | 0.957 (0.92, 0.99) | 0.974 (0.94, 1.00) |
| Risk (negative) | 0.099 (0.03, 0.18) | 0.159 (0.06, 0.28) | 0.102 (0.03, 0.19) | 0.154 (0.05, 0.28) |

The incremental contribution of genetic risk (PHS) differed by outcome. For early diagnosis of dementia, adding PHS to the multimodal framework produced only modest improvement beyond cognitive and imaging information alone. For AD differential diagnosis, by contrast, PHS provided a meaningful gain in discrimination, contributing to the strongest performance of the full multimodal model (AUC 0.91 in females and 0.83 in males) (Figure 1B), consistent with the established etiological specificity of genetic risk for AD.

Model discrimination was consistently higher in females across both outcomes and most predictor sets, with differences on the order of several AUC points. This pattern indicates potential sex-related differences in disease expression, biomarker signal-to-noise ratio, or measurement characteristics, and highlights the importance of sex-stratified validation when developing precision diagnostic tools for dementia and AD.

To confirm that the discrimination observed for early diagnosis of dementia was not driven by the inclusion of more advanced cases, we repeated the validation in subsets restricted to CDR ≤ 1 and CDR ≤ 0.5. Discrimination was preserved at CDR ≤ 1 (AUC ≈ 0.86 in both sexes) and remained high at CDR ≤ 0.5 (AUC ≈ 0.79 in males, AUC ≈ 0.82 in females), with sensitivity remaining high across all early-stage subsets (range: 0.91 to 0.96); full results are reported in Supplementary Table 1 and Supplementary Figures 4-5.

### Individual patient reports

Figure 2 exemplifies individualized patient reports generated for each outcome to translate model outputs into a clinically actionable format. The main panel shows the distribution of the combined model score in the reference data, plotted separately for cases and controls, with the x-axis partitioned into negative, inconclusive, and positive regions. Larger areas under a group’s density curve indicate that the corresponding group was more common in the reference data.

**Fig 2.**
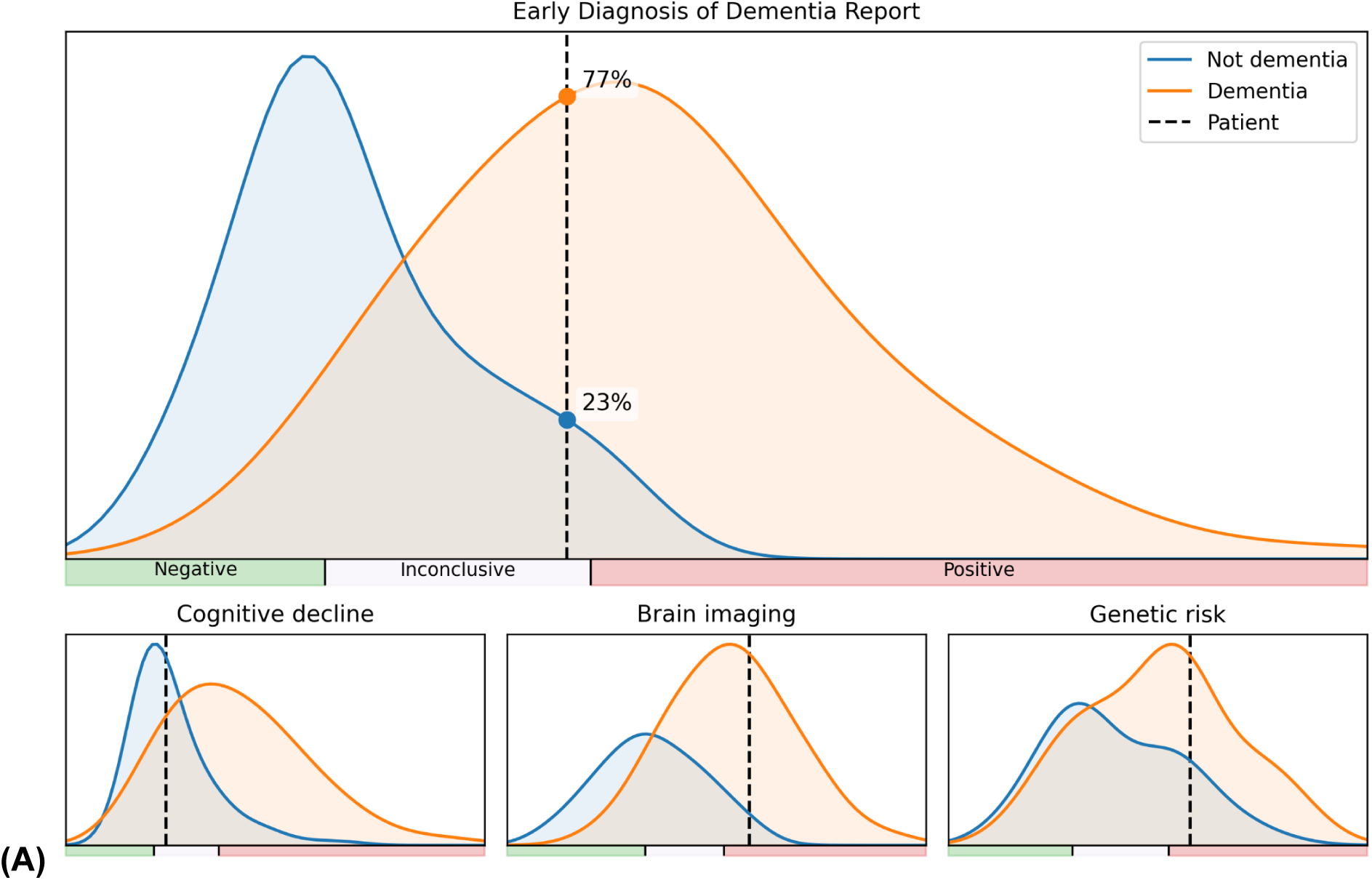
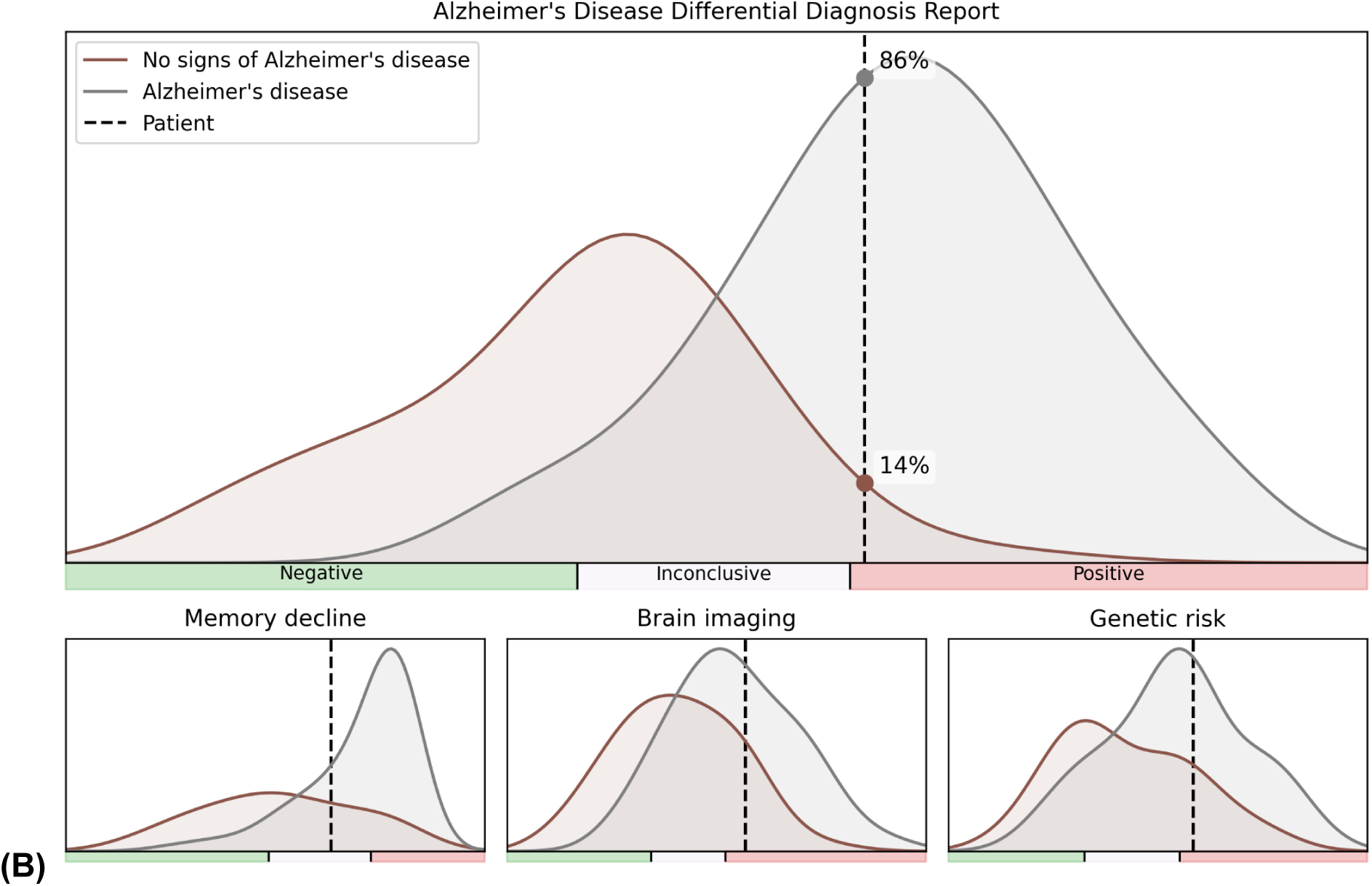
Prototype of individual patient reports for (A) early diagnosis of dementia and (B) differential diagnosis of Alzheimer’s disease. Each report presents the patient’s combined model score relative to the reference distribution of cases and controls, with the score axis divided into negative, inconclusive, and positive regions and the patient’s own score marked by a vertical dashed line. The three accompanying panels show the cognitive, imaging, and genetic component scores, with the patient’s value indicated on each for comparison with the reference population.

The patient’s own score is marked by a vertical dashed line, with point markers denoting estimated group membership probability at that score. Three component panels display the corresponding cognitive, imaging, and genetic component scores, with the patient’s value again marked to support intuitive comparison with the reference population.

## Discussion

In this study, we showed that the multimodal integration of cognitive assessments, neuroimaging, and polygenic data in the MHS-RWD tool yields strong discrimination for both early diagnosis of dementia and the differential diagnosis of AD in a real-world clinical cohort. The tool relies on data from routine clinical care including genetics. The complete multimodal models consistently outperformed individual predictors.

The two quantitative hazard components contributed to the models in distinct and complementary ways. The IHS originates from a model trained in ADNI, which is enriched for AD through its amnestic-MCI recruitment [24]. However, autopsy data indicate that ADNI cases are rarely pure AD pathologically, instead they span a range of other etiologies that frequently co-occur with AD [29,30]. The higher AUC of the IHS for early diagnosis of dementia than for AD differential diagnosis may indicate that the imaging signal learned in this setting captured a broad pattern of neurodegeneration rather than an AD-specific one. The PHS, by contrast, was developed using clinically diagnosed AD cases and therefore captures an AD-specific genetic liability. It contributed to the early diagnosis of dementia model despite mainly reflecting AD-specific risk, though its contribution was more modest than in the AD differential diagnosis model.

The sensitivity and specificity achieved by the MHS-RWD models in a real-world memory-clinic setting, particularly among female patients, were comparable to results obtained of emerging blood-based biomarkers for AD pathology in selected research cohorts. For example, plasma phosphorylated tau-217 (p-tau217), which has been validated against amyloid-PET or cerebrospinal fluid biomarkers, has shown ROC thresholds achieving 92–96% sensitivity and specificity [31–34]. Our MHS-RWD models for AD differential diagnosis achieved AUCs of 0.91 in females and 0.83 in males in a real-world cohort. These values approach the range of discrimination reported for p-tau217 in detecting AD pathology, and show the value of integrating cognitive, imaging, and genetic information for disease outcomes. It is important to note that p-tau217 assays are typically evaluated against biomarker-defined AD pathology, whereas our models predict clinical diagnostic outcomes in memory-clinic populations, which may reflect broader phenotypic variation and contribute to differences in sensitivity/specificity profiles. While direct head-to-head comparisons are limited by differences in reference standards and clinical endpoints, the performance of the MHS-RWD models supports their utility for both early diagnosis and etiological discrimination of dementia in clinical settings.

The rising incidence of AD globally calls for better and more cost-efficient tools for early diagnosis, with primary care expected to play an increasingly central role [2,3,8,35]. The MHS-RWD tool works well in real-world settings. It also seems to have a potential for deployment in primary care settings, where GPs can obtain the relevant assessments, and the MHS-RWD can integrate the different quantitative measures into a clinically useful early and differential diagnosis clinical decision support tool.

While the integration of genetic data in the MHS-RWD tool offers clear diagnostic utility, the use of genetic testing in dementia raises ethical considerations, particularly around risk disclosure, and downstream consequences for patients and their families. Genetic testing is acceptable when it enables clinically actionable decisions, a criterion that is increasingly met with the emergence of disease-modifying therapies for AD that benefit from early identification of individuals at elevated risk. Further demand for genotyping comes from evidence that certain treatments, including anti-amyloid monoclonal antibodies, may be contraindicated, restricted, or require intensified monitoring in individuals with high genetic risk, most notably *APOE* ε4 homozygotes [36,37]. It is a dilemma that people at greatest genetic risk may also face fewer therapeutic options and greater psychosocial and familial implications of genetic risk information.

To facilitate the use of MHS-RWD in clinical practice, patient level risk reports summarize multimodal risk estimates in an accessible visual format. The reports show the individual’s overall risk score relative to the distribution observed among cases and controls in the reference cohort, jointly with separate visualizations for cognitive, imaging, and genetic components, providing a sense of where the patient falls on the diagnostic spectrum. This transparency allows clinicians to see beyond a single, compound risk score and assess the domains contributing to the model’s outcome separately, aiming to support independent clinical reasoning without requiring direct interpretation of the underlying imaging or genetic data.

A limitation of our study is that the discrimination estimates reported here reflect evaluation against clinical diagnoses made in specialist memory clinics, not against neuropathological or biomarker-confirmed ground truth. Clinical diagnoses are subject to misclassification, and this diagnostic uncertainty may affect the observed model performance. At the same time, specialist memory-clinic diagnoses represent the highest-quality labels realistically available in large-scale clinical cohorts.

Another limitation of our study is that our sample was obtained from memory clinic cohorts, which represents a population that may differ from primary care patients in terms of age, disease severity, prevalence, and referral patterns. As a preliminary feasibility assessment, pilot testing of the MHS-RWD tool in primary care settings is currently ongoing. Moreover, our reliance on Norwegian samples limits the generalizability of these findings to populations with different ancestries and healthcare settings.

To conclude, integration of cognitive, neuroimaging, and polygenic data from routine clinical assessment in the MHS-RWD tool yields strong discrimination for both early diagnosis of dementia and AD differential diagnosis in a real-world clinical setting. Combined with the patient-level risk reports, the tool could support etiologically informed dementia assessment in primary care while reducing reliance on specialist health care.

## Data availability

Individual level data from the NorCog may be made available to researchers upon reasonable request, subject to approval by the relevant regulatory and governance bodies. Information on access procedures is provided in the NorCog cohort profile [20].

## Code availability

The codes for this study will be made available on https://github.com/ttfiliz/agecare_mhsrwd upon publication.

## Ethics declaration

Ethical approval for this study was granted by the Regional Committees for Medical and Health Research Ethics in Norway (REK 2013/2283-1 and REK 2014/631). The participants provided informed consent before inclusion. Access to and use of data and biological samples from the Norwegian Registry of Persons Assessed for Cognitive Symptoms is governed by regulations including the General Data Protection Regulation, the Health Register Act, the Health Research Act and the Register Regulations. NorCog requires that all projects using its data obtain approval from the Regional Committees for Medical and Health Research Ethics and the NorCog Steering Committee.

## Acknowledgements

The authors thank all patients and informants for providing information. We are grateful for the efforts made by the reporting outpatient clinics and the steering committee for the Norwegian Registry of Persons Assessed for Cognitive Symptoms. Funding was provided by the Research Council of Norway (RCN) #344121. This work also used the TSD (Tjenester for Sensitive Data) facilities, owned by the University of Oslo, operated and developed by the TSD service group at the University of Oslo, IT-Department (USIT)

