## Supplementary Information for "An Integrative Multimodal Model for Early Diagnosis of Dementia and Differential Diagnosis of Alzheimer’s Disease Using Neuroimaging, Polygenic Risk, and Cognitive Assessments"

Supplementary Table 1. **Sex-stratified discrimination and score-band accuracy for models developed for early diagnosis of dementia (A) and differential diagnosis of Alzheimer's disease (B), reported for the full validation set and for subsets constrained to CDR ≤ 1 and CDR ≤ 0.5.** Sensitivity and specificity are reported conditional on receiving a definitive classification. 95% confidence intervals are denoted in parentheses.

(A)

| Metric | Main: Male | Main: Female | CDR ≤ 1: Male | CDR ≤ 1: Female | CDR ≤ 0.5: Male | CDR ≤ 0.5: Female |
|---|---|---|---|---|---|---|
| N | 353 | 292 | 152 | 128 | 108 | 85 |
| Sensitivity | 0.946 (0.905, 0.981) | 0.949 (0.905, 0.983) | 0.955 (0.900, 1.000) | 0.944 (0.877, 1.000) | 0.906 (0.793, 1.000) | 0.955 (0.850, 1.000) |
| Specificity | 0.780 (0.684, 0.867) | 0.914 (0.836, 0.980) | 0.742 (0.581, 0.889) | 0.850 (0.682, 1.000) | 0.733 (0.565, 0.889) | 0.842 (0.667, 1.000) |
| Coverage | 0.601 (0.547, 0.652) | 0.603 (0.545, 0.658) | 0.645 (0.566, 0.717) | 0.578 (0.492, 0.664) | 0.574 (0.481, 0.667) | 0.482 (0.376, 0.588) |
| AUC | 0.843 (0.798, 0.883) | 0.893 (0.855, 0.929) | 0.856 (0.796, 0.909) | 0.858 (0.788, 0.920) | 0.788 (0.700, 0.867) | 0.820 (0.725, 0.905) |

(B)

| Metric | Main: Male | Main: Female | CDR ≤ 1: Male | CDR ≤ 1: Female | CDR ≤ 0.5: Male | CDR ≤ 0.5: Female |
|---|---|---|---|---|---|---|
| N | 219 | 193 | 96 | 89 | 65 | 54 |
| Sensitivity | 0.908 (0.839, 0.971) | 0.926 (0.861, 0.976) | 0.947 (0.865, 1.000) | 0.939 (0.848, 1.000) | 0.909 (0.783, 1.000) | 0.929 (0.769, 1.000) |
| Specificity | 0.661 (0.526, 0.780) | 0.943 (0.857, 1.000) | 0.818 (0.643, 0.955) | 1.000 (1.000, 1.000) | 0.875 (0.692, 1.000) | 1.000 (1.000, 1.000) |
| Coverage | 0.603 (0.534, 0.667) | 0.601 (0.528, 0.668) | 0.625 (0.521, 0.719) | 0.551 (0.449, 0.652) | 0.585 (0.462, 0.708) | 0.519 (0.389, 0.648) |
| AUC | 0.828 (0.771, 0.879) | 0.906 (0.860, 0.947) | 0.881 (0.810, 0.941) | 0.918 (0.855, 0.966) | 0.867 (0.761, 0.958) | 0.949 (0.884, 0.993) |

Supplementary Table 2. **Distribution of Clinical Dementia Rating (CDR) global scores in the validation set for early diagnosis of dementia (A) and differential diagnosis of Alzheimer's disease (B), stratified by sex.**

(A)

| CDR | Male N | Female N |
|---|---|---|
| 0 | 12 | 14 |
| 0.5 | 96 | 71 |
| 1 | 44 | 43 |
| 2 | 10 | 13 |
| 3 | 0 | 2 |
| Missing | 191 | 149 |

(B)

| CDR | Male N | Female N |
|---|---|---|
| 0 | 5 | 5 |
| 0.5 | 60 | 49 |
| 1 | 31 | 35 |
| 2 | 8 | 9 |
| 3 | 0 | 2 |
| Missing | 115 | 93 |

**Supplementary Figure 1.** Reference distributions for early diagnosis of dementia for female (panel A) and male (panel B) participants.

(A)

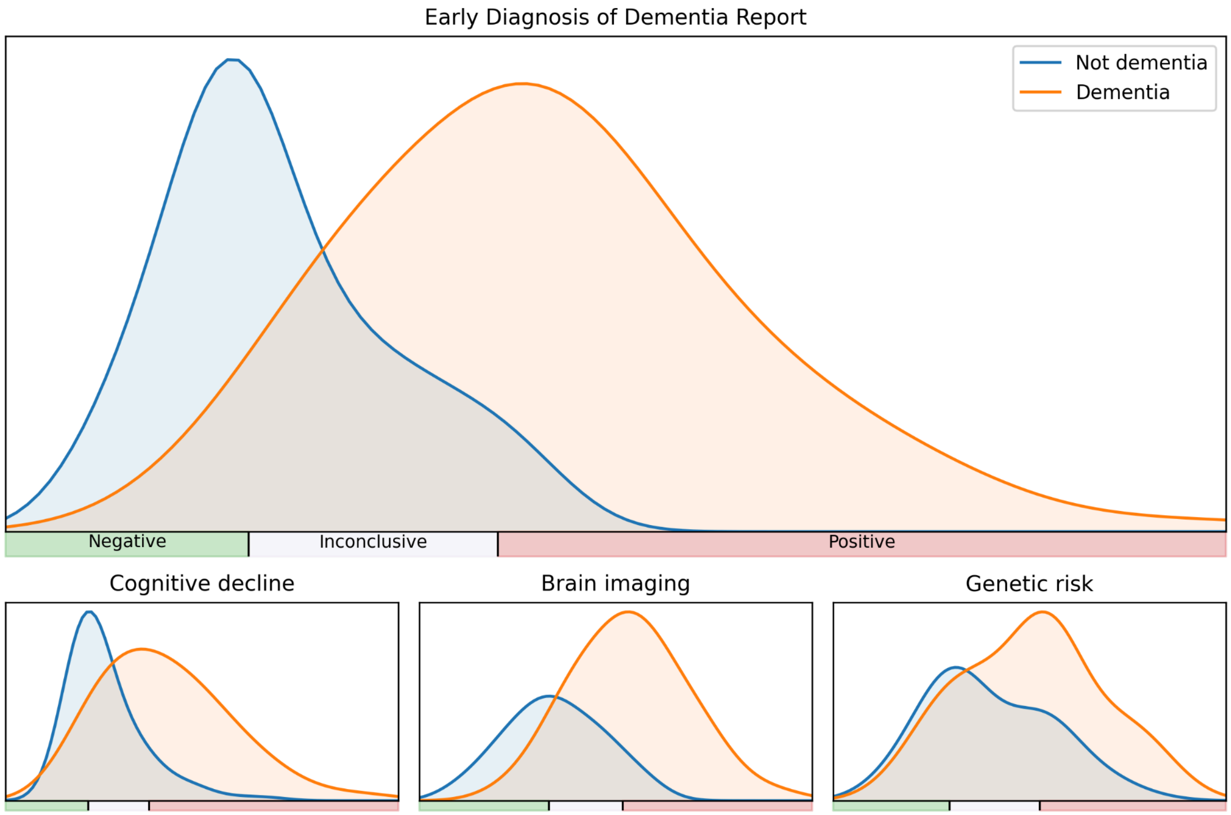

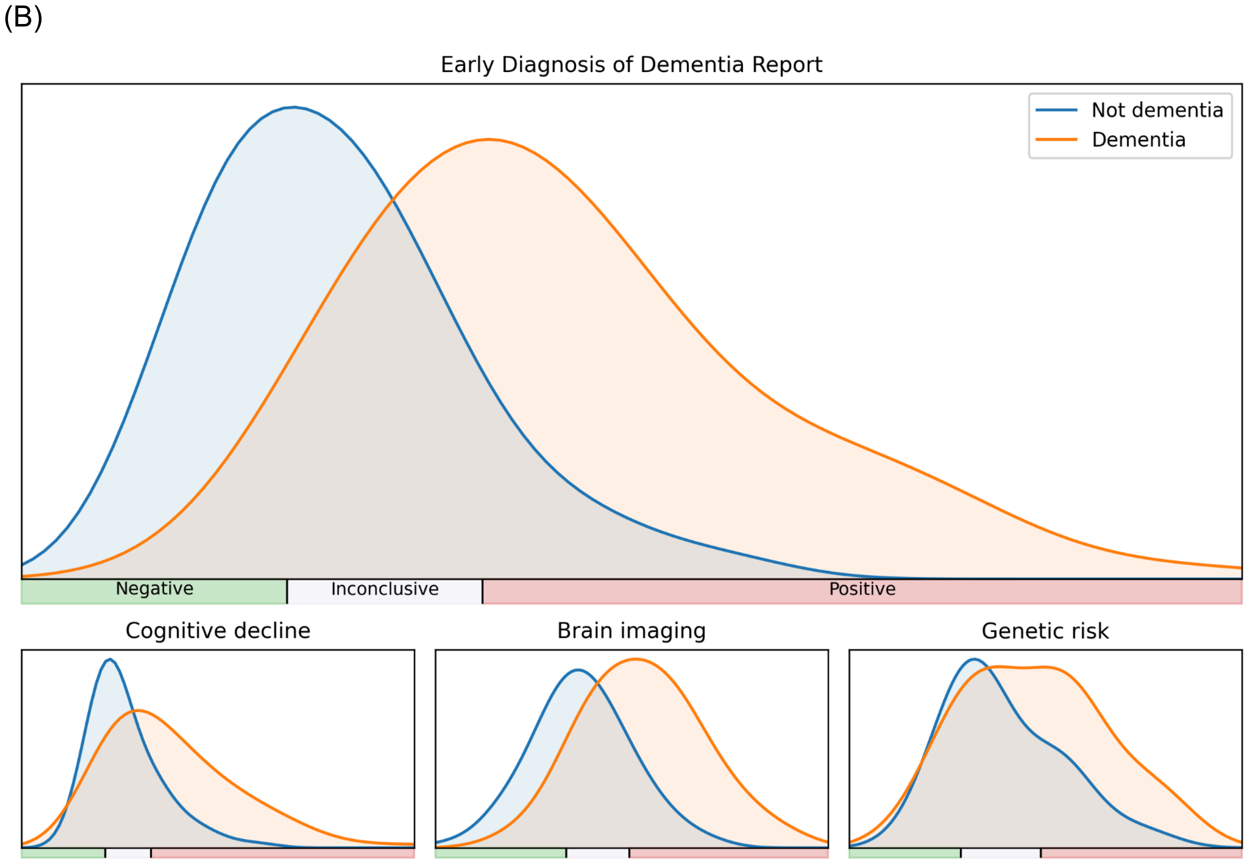
(B)
Early Diagnosis of Dementia Report
Not dementia
Dementia
Negative
Inconclusive
Positive
Cognitive decline
Brain imaging
Genetic risk

**Supplementary Figure 2.** Reference distributions for AD differential diagnosis for female (panel A) and male (panel B) participants.

(A)

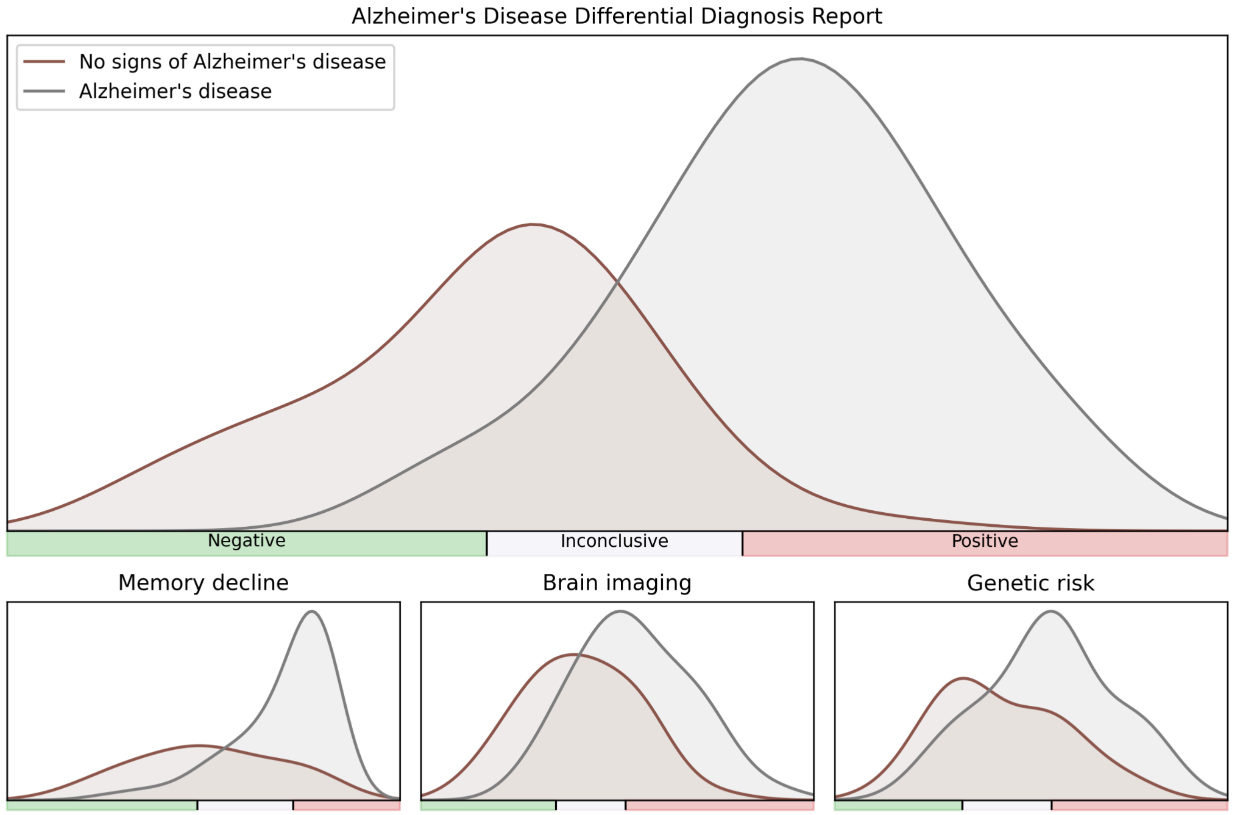

(B)

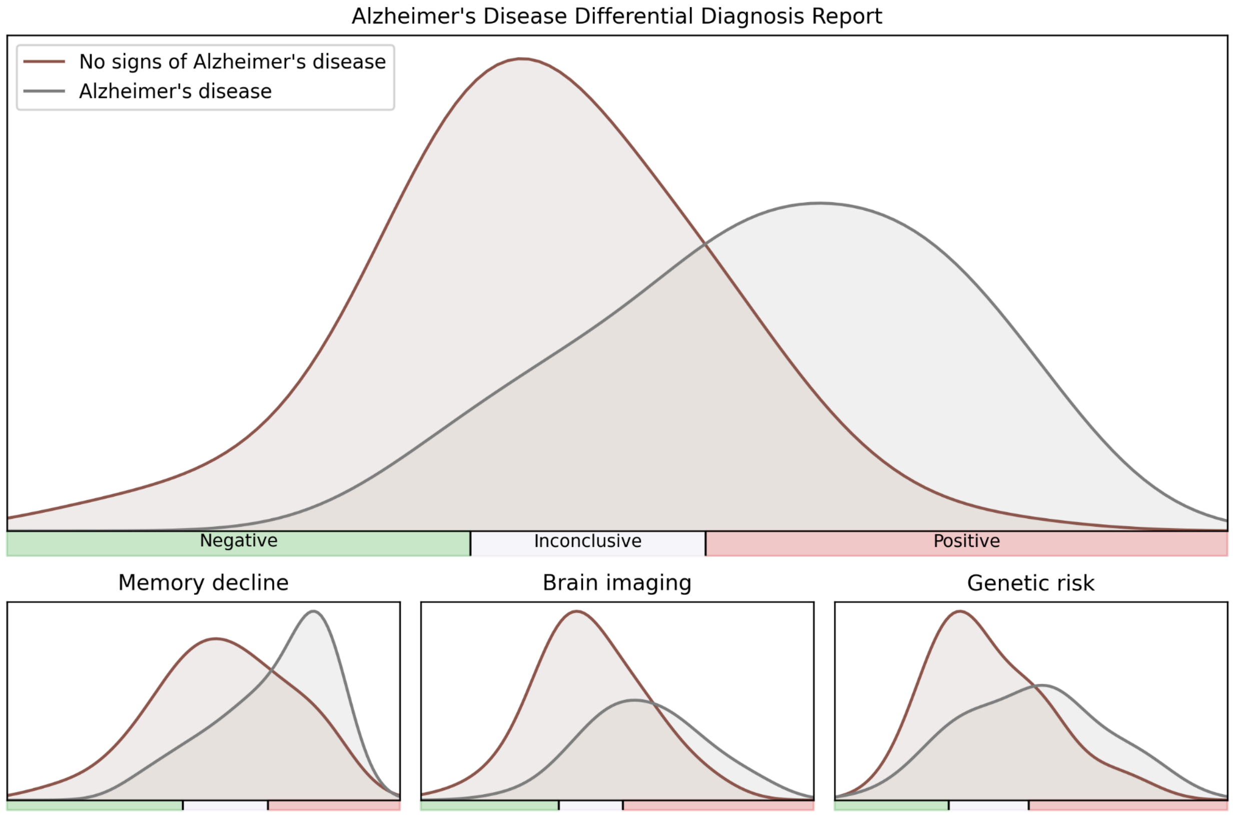
Alzheimer's Disease Differential Diagnosis Report
No signs of Alzheimer's disease
Alzheimer's disease
Negative
Inconclusive
Positive
Memory decline
Brain imaging
Genetic risk

**Supplementary Figure 3.** Flowchart of cohort selection and outcome-specific analytic samples for the full MHS-RWD models. Flowchart showing cohort selection from the St. Olav's Hospital and Oslo University Hospital (OUS) memory-clinic cohorts for the two full-model analyses: differential diagnosis of Alzheimer's disease (AD) and early diagnosis of dementia. Numbers are shown overall and, where applicable, stratified by cohort and sex. N, sample size; M, male; F, female; SCI, subjective cognitive impairment; MHS-RWD, Multimodal Hazard Score for Real-World Data.

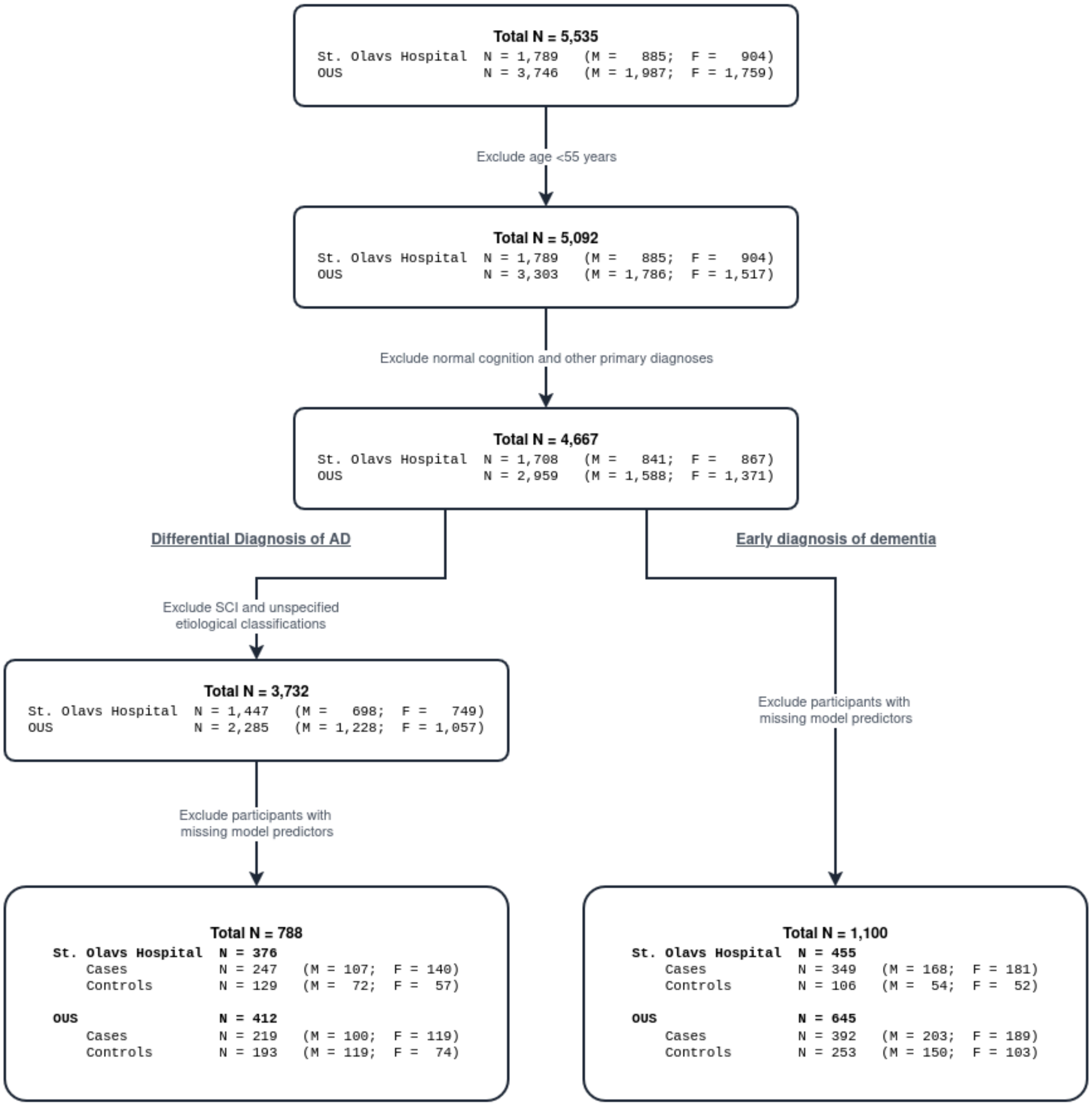

**Supplementary Figure 4. Accuracy of the models for early diagnosis of dementia and differential diagnosis when the validation set is constrained to participants with CDR ≤ 1.** The figure presents sex-stratified model performance and risk stratification for the two outcomes. Panel (A) shows results for early diagnosis of dementia; panel (B) shows results for Alzheimer's disease differential diagnosis. In each panel, receiver operating characteristic (ROC) curves are displayed separately for females and males, comparing a baseline age-only model with MHS-RWD; the corresponding area under the curve (AUC) values are reported in the legends. Star and dot markers on the ROC curves indicate predefined percentile-based thresholds used to define negative, inconclusive (grey zone), and positive score regions. To the right of each ROC pair, bar plots summarize the sex-stratified probability of the clinical outcome within these three score categories.

(A)

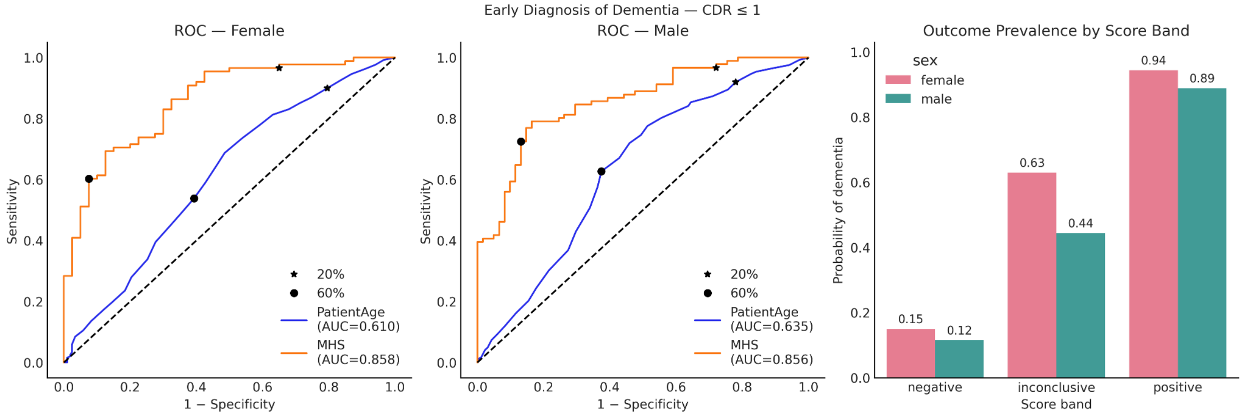


(B)

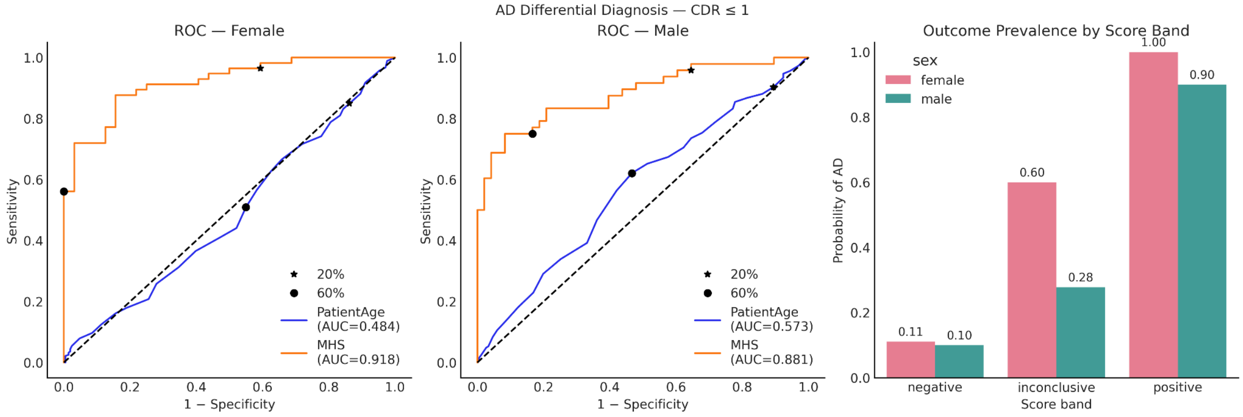

**Supplementary Figure 5. Accuracy of the models for early diagnosis of dementia and differential diagnosis when the validation set is constrained to participants with CDR ≤ 0.5.**

The figure presents sex-stratified model performance and risk stratification for the two outcomes. Panel (A) shows results for early diagnosis of dementia; panel (B) shows results for Alzheimer's disease differential diagnosis. In each panel, receiver operating characteristic (ROC) curves are displayed separately for females and males, comparing a baseline age-only model with MHS-RWD; the corresponding area under the curve (AUC) values are reported in the legends. Star and dot markers on the ROC curves indicate predefined percentile-based thresholds used to define negative, inconclusive (grey zone), and positive score regions. To the right of each ROC pair, bar plots summarize the sex-stratified probability of the clinical outcome within these three score categories.

(A)

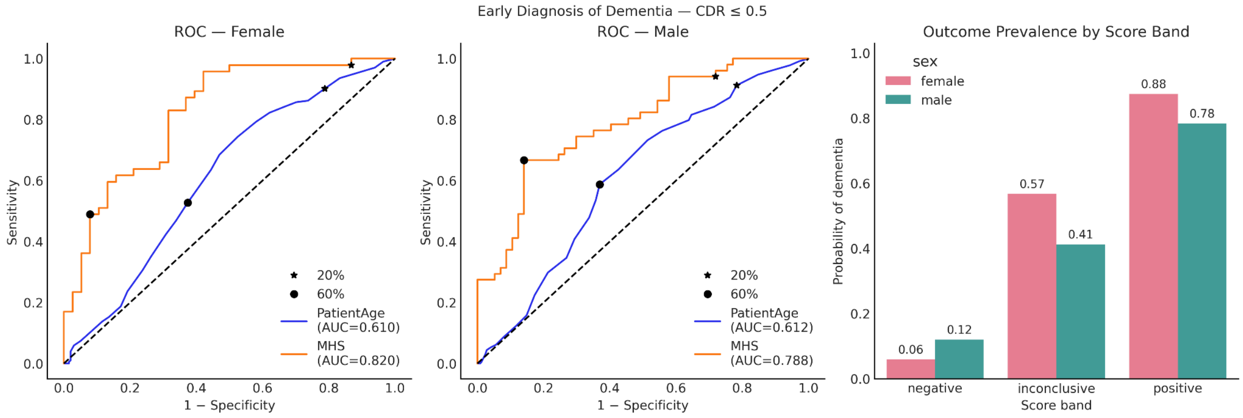


(B)

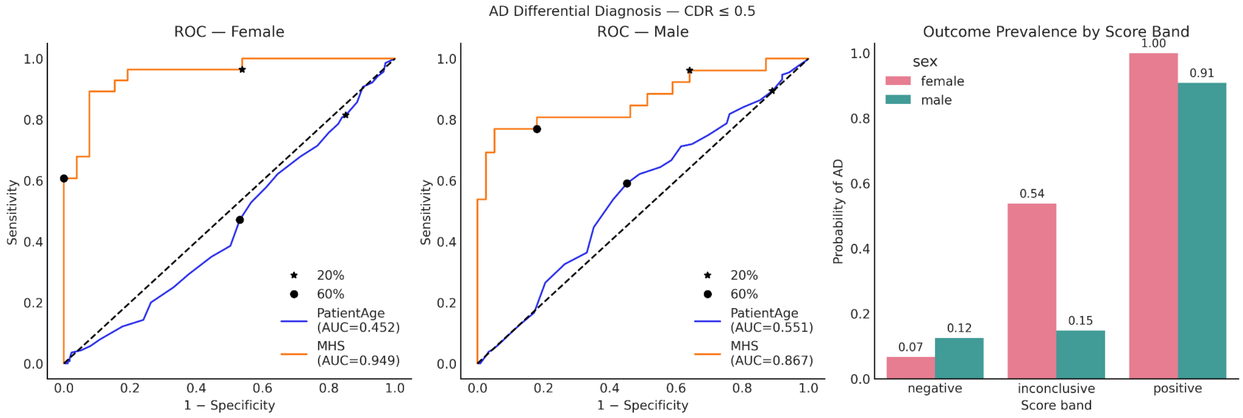
